# REPROGRAM: REsilience PROmotion with GeRoprotectors: AssessMent of biological effect. Rationale and protocol for a trial of biological effect

**DOI:** 10.64898/2026.03.19.26348863

**Authors:** Daisy Wilson, Animesh Acharjee, Niharika A Duggal, Jose R Hombrebueno, Simon W Jones, Jonathan Lewis, João Pedro de Magalhães, Yessica Martinez-Serrato, Ali Mazaheri, Helen M. McGettrick, Sudip Mondal, Amy J Naylor, Aline Nixon, Thomas Nicholson, Judith Partridge, Thomas Pinkney, Nicholas J W Rattray, Claire Steves, Kristina Tomkova, Carly Welch, Thomas Jackson

## Abstract

**Background:** Ageing is associated with reduced resilience to physiological stressors such as infection and surgery. This reduced resilience is believed to be underpinned by the hallmarks of ageing, the key biological mechanisms driving the aged phenotype. Geroprotectors are drugs that are proposed to slow down the ageing process and promote longevity and healthspan. Despite this, mechanistic studies in healthy older adults are lacking.

**Methods and Analysis:** This trial will test the hypothesis that geroprotectors targeted towards biological mechanisms associated with poor resilience can reverse these pathways within a three-week period. Three geroprotectors with a good safety profile in older adults and evidence of effect on the hallmarks of ageing will be administered to 60 (30 female; 30 male) adults 70+. Participants will be randomised to one of three arms (Metformin MR 1500mg, Fisetin 100mg or Spermidine 15mg). Participants will be extensively clinically characterised at baseline. Blood, abdominal adipose tissue and stool samples will be taken at baseline and following the three-week intervention. The primary research question will answer whether a three-week course of Metformin, Spermidine, or Fisetin reduce the number of senescent cells as measured by SA-β-GAL in adipose biopsies in healthy older volunteers. Additionally, there will be assessment of the effect of the geroprotectors on other hallmarks of ageing, including autophagy, immunosenescence, chronic inflammation, dysregulated mTOR signalling, epigenetic age, DNA damage, dysregulated metabolism, stem cell exhaustion and microbial composition.

**Ethics and Dissemination:** Ethical approval is in place (24/LO/0549). The main trial report and any sub-studies will be published in high impact peer-reviewed gerontology journals, presented at academic conferences and through a series of public engagement events. Participants enrolled in the study will be informed of the results by a written summary.

**Trial Registration:** REPROGRAM was registered with ISRCTN on 10/09/24. ISRCTN47919839. Available at https://www.isrctn.com/search?q=47919839.

**Trial Registration Data Set:** Table 1
Trial Registration Data Set

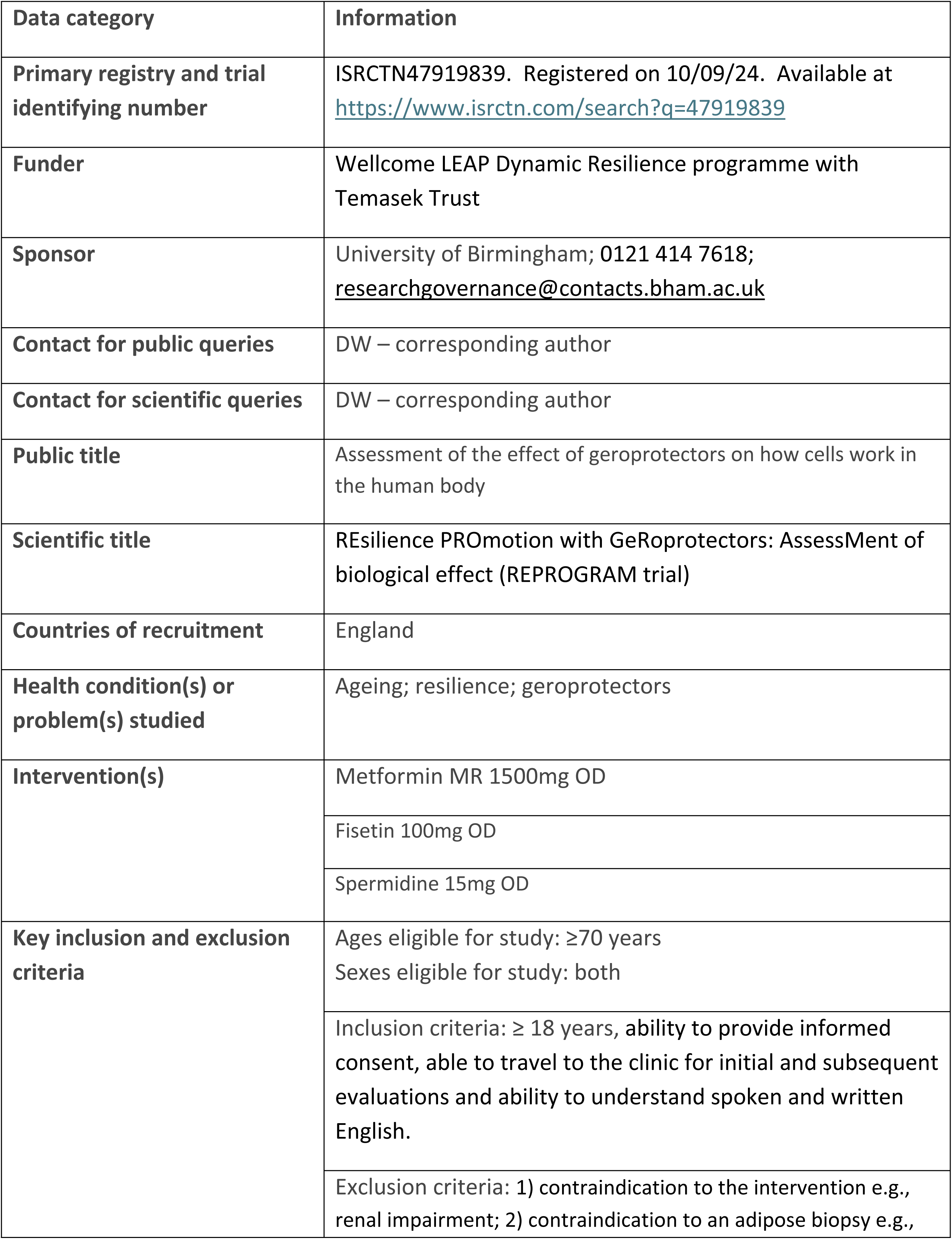

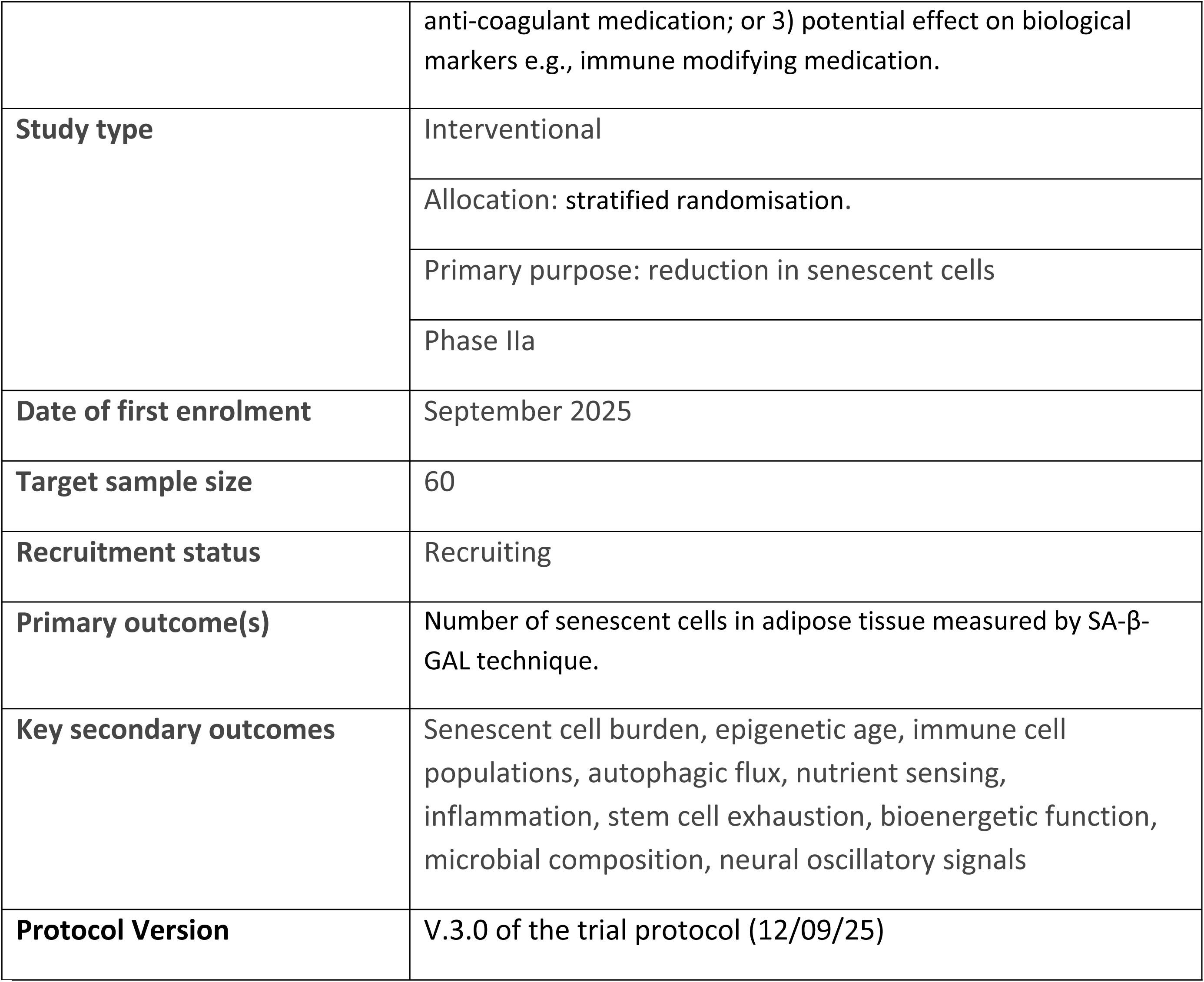

## Introduction

### Background and Rationale

Ageing is associated with reduced resilience, hindering the ability to maintain homeostatic balance when confronted with stressors such as infections, falls, or surgery(1). Advances in ageing biology have enabled the description of key biological mechanisms driving the aged phenotype, the hallmarks of ageing(2), including: reduced mitochondrial fitness, telomere shortening, reduced autophagy, accumulation of senescent cells, microbial dysbiosis and increased systemic inflammation. Importantly, targeting these ageing processes extends health-span in animal models(3, 4).

Geroprotectors are drugs that can affect underlying ageing processes rather than a single symptom or disease. However, there is an absence of robust pharmacodynamic and pharmacokinetic data on their ability to modify the proposed targets *in vivo* in humans. Additionally, it is unclear which patients should be treated and where in their life-course the drugs will have the optimum effect(5). This is essential to establish to enable a stratified medicine approach where interventions are carefully targeted to successfully prevent and/or treat clinical manifestations of impaired resilience in older people in ways that ensure treatment is most likely to benefit them and not cause harm.

This study aims to investigate three different agents, Metformin, Fisetin and Spermidine, and determine their ability to modify biological pathways in vivo in humans. These agents have evidence of effect on the hallmarks of ageing and additionally good safety profiles for use in older adult cohorts (6–10) (Supplementary Materials for complete side effect profiles of each agent). Moreover, these agents individually target different hallmarks of ageing, thus maximising the opportunity to identify biological mechanisms of ageing sensitive to manipulation from geroprotectors.

Metformin, an oral hypoglycaemic agent, primarily acts on nutrient sensing pathways: decreasing insulin and IGF-1 levels (11), increasing insulin sensitivity (11), acting as AMPK activator (12) and inhibiting mTOR (13, 14). It also appears to slow biological ageing through acting on other hallmarks including autophagy (15–20), cellular senescence (2, 21–25), inflammation (13–15, 26–29), telomere attrition (11, 12, 30, 31), stem cell exhaustion (17, 32–36), DNA damage (37–43) and oxidative stress (41, 44–46)).

Studies using animal models to investigate the effect of Metformin on health and life-span are inconclusive with reports of both positive results in mice (47, 48) and C.elegans (12) and negative or no difference results in rats (49) and Drosophilia (50). A meta-analysis of Metformin in healthy mice and C. elegans found heterogenous effects but concluded it was not associated with overall life-span prolonging effects (51).

Meta-analysis of 53 human clinical cohort studies found lower mortality, reduced incidence of cancer and cardiovascular disease in individuals taking Metformin for diabetes (52).

Metformin improved gait speed in pre-frail, non-diabetic individuals (52–54) and reduced the rate of decline in gait speed in diabetic women (55). The evidence is currently inconclusive as other human studies have found a negative impact of Metformin on hypertrophic responses to resistance training (56) and no improvement in gait speed or physical performance with Metformin (56, 57).

Fisetin targets cellular senescence through the senescent-cell anti-apoptotic pathways (58, 59) reducing the number of senescent cells in animal and tissue models (60, 61). It is a plant flavanol, found in edible fruits and vegetables such as strawberries, apples and persimmons(62). In culture models Fisetin decreased the viability and number of senescent cells (59, 61, 63–65). In vivo, Fisetin reduced senescence and inflammation in both mice (66–68) and aged sheep (69). Fisetin administered to aged wild-type mice at 100mg/kg dose has been demonstrated to extend both life and health-span (58). Clinical trials of 100mg of fisetin once a day in humans have demonstrated an anti-inflammatory effect, reduction in IL-8 in patients with cancer receiving chemotherapy supplemented with fisetin (70), reduction in MMP-2, MMP-9 and CRP in patients suffering an acute ischaemic stroke who received fisetin alongside recombinant tissue plasminogen activator (71) and, reduction in senescence-associated secretory phenotype in healthy participants (72, 73).

Spermidine, a natural polyamine, extends life-span through autophagy by inhibiting EIA-associated protein p300 and preventing acetylation of cytosolic autophagy related proteins (74, 75). Spermidine is found in many food products, including dry soybeans, chicken liver and blue cheese. It is an essential metabolite that exhibits age-dependent decline in a number of models including humans (76). Spermidine increased cell survival of human PBMCs in culture (77). In yeast, C. elegans, flies and mice Spermidine extended life-span (77, 78) and in rat models Spermidine increased life and health-span (79). Epidemiological studies have demonstrated that diets high in Spermidine are associated with lower mortality(80). A review of three trials administering Spermidine to patients with cognitive dysfunction found conflicting results with two reporting improvement in cognitive function (81, 82) and one reporting no difference (83).

The current lack of mechanistic studies of the effects of these agents on the biological hallmarks of ageing in healthy older adult cohorts alongside the conflicting evidence within the literature highlights the need for the research investigating underlying biological pathways alongside clinical outcomes. This will underpin future carefully designed trials in frail older adult cohorts with greater specificity for targeting improved health-span.

The dosing strategy for this trial has been determined using the available literature. All previous clinical trials of fisetin have used 100mg (70, 71, 73, 84); this was originally adopted, based on the FDA model of conversion from an *in vivo* dose to the safe human equivalent (70). Previous clinical trials of Metformin have used between 1500mg to 2000mg ((56, 85–89). A dose of 1500mg was selected as it reflects a typical dose used in diabetes and enables safe, gradual up-titration within the short trial time-frame. A dose of 15mg of Spermidine was selected after discussion with experts and review of the seminal paper by Eisenberg et al examining Spermidine across several models (77). The previously reported clinical trials used much lower doses (0.9-3.3mg) (81–83) but commercially available supplements are typically between 7-10mg/day.

This trial is running in conjunction with REBOUND a prospective cohort study investigating the fundamental mechanisms of accelerated ageing and impaired resilience following cancer surgery and treatment (90). REPROGRAM and REBOUND are both funded by a Wellcome LEAP Dynamic Resilience Award and their study design and experimental analysis is complementary. The overarching aim of both studies is to determine whether biological hallmarks associated with the worsening of physical and cognitive function identified in REBOUND are ameliorable to improvement with a geroprotector.

### Objectives

This trial aims to determine whether targeted interventions towards underlying biological mechanisms associated with poor resilience can reverse biological pathways within a three-week period.

#### Primary Research Question

- Does a three-week course of Metformin MR, Spermidine, or Fisetin at intervention dose reduce the number of senescent cells as measured by SA-β-GAL in adipose biopsies in healthy older volunteers?

#### Secondary Research Questions

- Does a three-week course of Metformin MR, Spermidine, or Fisetin at intervention dose: 1) Reduce senescent cell burden in adipose tissue as measured by transcriptomics using SenMayo analysis, 2) Reduce epigenetic age (DNA methylation) from whole blood and adipose tissue, 3) Alter immune cell frequencies and features of immunosenescence, 4) Improve autophagic flux as measured by flow cytometry, 5) Improve nutrient sensing as measured by mTOR activation, 6) Improve mitochondrial function as measured by Seahorse, 7) Reduce inflammation as measured by ELISA and multiplex technology, 8) Improve stem cell exhaustion as measured in adipose derived stem cells, 9) Change microbial composition and functional potential of stool using metagenomics, 10) Change in neural oscillatory signatures measured by EEG

#### Trial design

This is a Phase 2a clinical trial on healthy volunteers which will be conducted at a single site, University Hospitals Birmingham NHS Foundation Trust. It is an experimental trial designed to mimic a surgical window of opportunity trial.

## Methods

This protocol was written with adherence to SPIRIT reporting guidelines (91). Recruitment started on 24/09/25 and will be complete by April 2026. Data collection will be completed by July 2026 and results are expected October 2026.

### Trial setting

Healthy volunteers will attend the Clinical Research Facility (CRF) at Queen Elizabeth Hospital Birmingham, UK for a total of three visits. Figure 1 outlines the visits to the CRF and the participant timeline.

**Figure 1.**
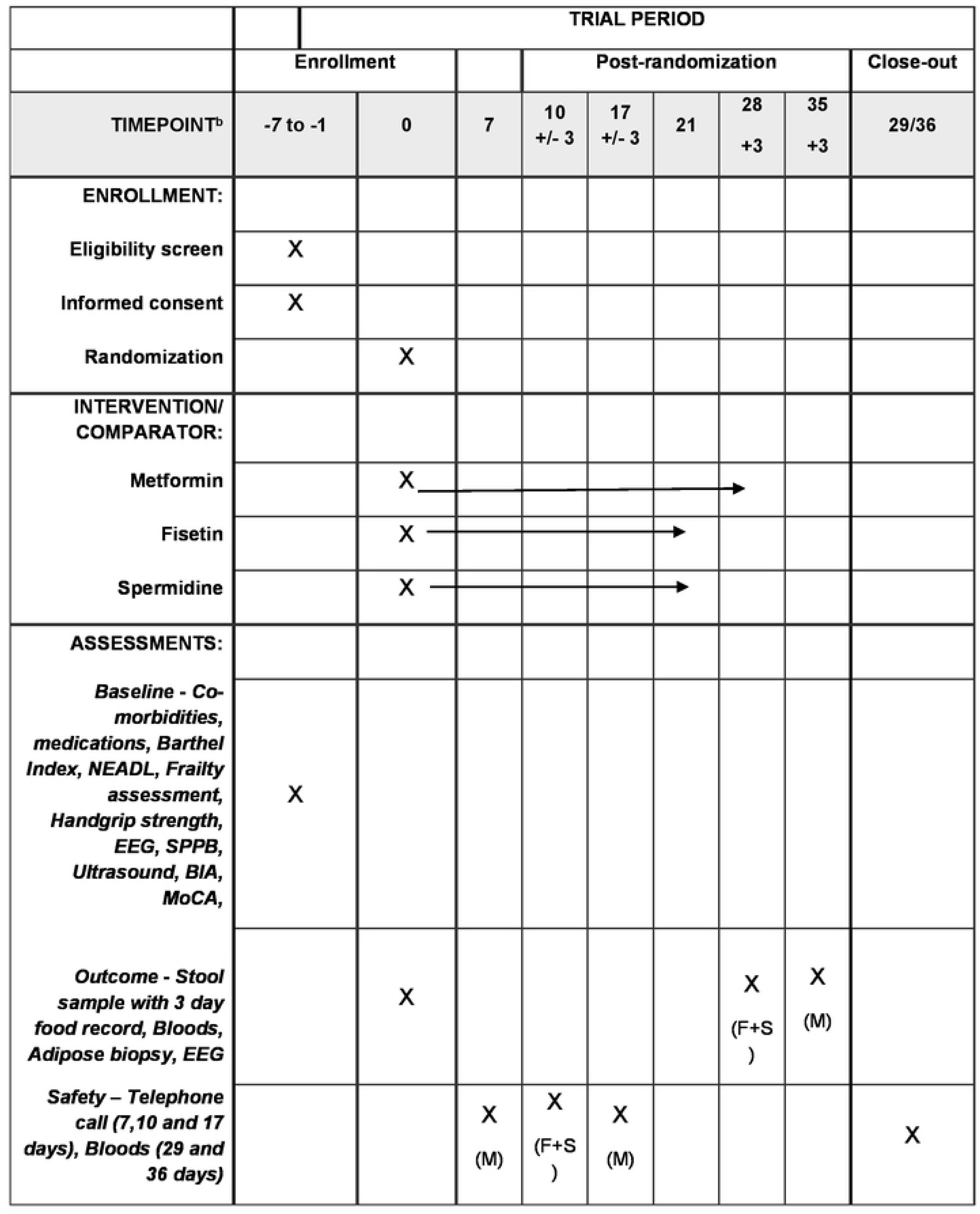
SPIRIT Figure - Participant timeline: Schedule of enrollment, interventions, and assessments.^a^ (92) Participant Timeline: The intervention is given for 3 weeks at trial dose. To reduce the risk of side effects Metformin is given for 1 week at lower dose prior to increasing to the trial dose. Therefore, there is an additional telephone call at Day 7 to discuss whether to increase the dose to trial dose and the mid-point telephone call and outcome assessment are 7 days later compared to Fisetin and Spermidine. In the figure M denotes Metformin and F and S denote Fisetin and Spermidine respectively.

In brief, following an expression of interest from a healthy volunteer they will be contacted by the trial team and asked to attend the CRF for the first visit. The first visit comprises of consent, clinical assessment and blood tests for safety checks and biological assessment.

The second visit, one week following the first visit, comprises of adipose biopsy, collection of stool sample, randomisation and distribution of the intervention. The participants taking Spermidine and Fisetin will all be contacted at home approximately ten days after starting the intervention to check on side effects. Participants taking Metformin MR (Modified Release) will be contacted one week after starting to discuss side effects and increase the dose to treatment dose. They will then be contacted at home approximately ten days after the dose increase to check on side effects. The third visit, one week after completing the intervention, comprises of adipose biopsy, collection of stool sample, blood tests for safety checks and biological assessment and assessment of adherence to intervention (pill count and diary review).

### Eligibility criteria

Inclusion criteria are: age ≥70 years, ability to provide informed consent, able to travel to the clinic for initial and subsequent evaluations and ability to understand spoken and written English.

The exclusion criteria are listed in full within Table 1 in the supplementary materials but are broadly split into three criteria, exclusion due to:

1. contraindication to the intervention e.g., renal impairment;
2. contraindication to an adipose biopsy e.g., anti-coagulant medication; or
3. potential effect on biological markers e.g., immune modifying medication.

Participants with coeliac disease will be excluded from taking spermidine as it is a wheat germ extract.

### Consent

Written consent will be taken by a trained member of the research team. Participants will be consented for blood and stool collection, and adipose biopsies. There will be specific optional consent for samples and anonymised research data to be used in future ethically approved research.

### Intervention

The justification for the choice and dose of Metformin MR, Spermidine and Fisetin has been established in the background section but in conclusion there is evidence of their efficacy in pre-clinical models alongside a good safety profile in older adults. There is no placebo arm as the trial is an investigation of biological endpoints. The three-week duration of intervention has been selected to mirror a surgical window of opportunity trial with the expectation that a successful agent could be deployed as a prehabilitation intervention.

Metformin MR will be given at a lower dose for one week prior to the three-weeks at treatment dose to reduce the risk of side effects.

There are three intervention arms – Metformin MR, Spermidine and Fisetin. All three geroprotectors will be taken orally with or just after food in the evening (See supplementary materials for details of suppliers and preparations).

#### BOX 1

Intervention Arms

1. Metformin MR – 500mg for one week (starting at a low dose to minimise the risk of side effects) increased to 1500mg the treatment dose for a further three weeks.
2. Spermidine – 15mg for three weeks.
3. Fisetin – 100mg for three weeks.

Investigators may discontinue the intervention in the event of side effects occurring that are not tolerated by the participant, or constitute a serious adverse reaction (SAR) or suspected unexpected SAR. The intervention will also be discontinued at a participants request or in the event of a new diagnosis that features on the exclusion criteria. Investigators may not reduce the dose.

Participants who discontinue the intervention and wish to remain in the trial will be followed up as per their allocated treatment intervention arm. For participants who discontinue the intervention but don’t withdraw from the trial the Investigator will consider the scientific importance of post-intervention blood samples, adipose biopsies, stool samples and EEG and balance this against the risks and burden of each assessment before recommending whether to continue with the full assessment or not.

To improve adherence to the interventions: a once-a-day dosing strategy has been employed, and Metformin MR has been selected rather than a non-modified release preparation of metformin. Metformin MR has a superior side effect profile to non-modified release metformin(93).

### Outcomes

This is a trial of the biological effect of the three interventions and the outcomes reflect this. The primary outcome is:

- Number of senescent cells in adipose tissue measured by SA-β-GAL technique after three weeks at intervention dose, adjusted for baseline number of senescent cells prior to the intervention.

The secondary outcomes will describe the effect of the three interventions on the change from baseline to after three weeks of intervention in:

1. senescent cell burden in adipose tissue as measured by transcriptomics using SenMayo analysis,
2. epigenetic age (DNA methylation) from whole blood and adipose tissue,
3. altered immune cell frequencies and features of immunosenescence to calculate Immunological age (IMM-AGE score), through spectral flow cytometry of peripheral blood mononuclear cells (PBMCs),
4. autophagic flux assessed by quantifying lysosomal LC3 accumulation following inhibition of lysosomal degradation, using flow cytometry
5. nutrient sensing assessed by quantifying phosphorylation of pS6K in immune cell subsets by flow cytometry as a downstream readout of mTOR signalling
6. cellular bioenergetic function, including mitochondrial oxidative phosphorylation and glycolytic metabolism, quantified using real-time flux-based respirometry
7. inflammation as measured by ELISA and multiplex technology,
8. stem cell exhaustion through the isolation and differentiation of adipose derived stem cells,
9. microbial composition and functional potential of stool using metagenomics.
10. neural oscillatory signatures measured by EEG

### Sample Size

The sample size calculation is based on measurements of SA-β-Gal from a senolytic trial in adults (94). The number of participants required at 80% power and a significance level of 0.05 to detect a mean difference of 5% with a standard deviation of 4 in SA-β-Gal marker is 9. To allow for drop out 10 participants of each sex will be recruited to each intervention (Metformin, Spermidine, or Fisetin), 20 per arm (60 in total). We are recruiting 10 participants of each sex for each intervention as there is some evidence of sex differences with women believed to be more prone to senescence (95).

### Recruitment

Volunteers will be approached via email through the Birmingham 1000 Elders database of research active healthy older people who have consented to be contacted about studies run through the University of Birmingham. We have used this approach successfully before when recruiting to a trial investigating the biological effect of a nutraceutical(96). Additionally, we will be approaching established community networks and utilising the NIHR funded ‘Be Part of Research Registry’.

## Assignment of interventions

### Allocation

The trial uses a stratified randomisation technique (blocked randomisation list) to ensure the ages of participants are evenly distributed across the interventions (97, 98).

Randomisation is completed by Investigator at Visit 2. In the event of inadequate recruitment, an open-label design may be adopted to enable effective completion of specific trial arms

### Blinding

The clinical team and participants will not be blinded to the intervention for safety and procedural reasons. The laboratory scientists will be blinded to the intervention during their experiments and analysis. Samples will be labelled with participant ID and they will have no access to corresponding clinical or intervention data.

## Data collection and management

### Assessment and collection of outcomes

Figure 1 describes the schedule of events that will be conducted as part of this trial. REPROGRAM is running alongside REBOUND, a cohort study investigating the fundamental mechanisms of accelerated ageing and impaired resilience following cancer surgery and treatment. The research procedures used to describe the participants for REPROGRAM are very similar to REBOUND and the justification for individual techniques has been discussed in the REBOUND protocol (90).

In brief, this trial depends upon accurate description of participants and correlation with biological outcomes. Clinical characterisation of the participants consists of:

1. Cognitive assessment (MoCA),
2. Muscle quantity and quality assessment (quadriceps ultrasound, BIA),
3. Muscle physical function assessment (hand grip strength, physical performance),
4. Frailty and sarcopenia diagnoses (Frailty phenotype, Clinical Frailty Scale and Sarcopenia informed by above measures and Nottingham Extended Activities of Daily Living).

### Data Management

Source data will be collected on paper CRFs and transferred to eCRFs on REDCap. REDCap is a secure encrypted data management software. Access to REDCap will be provided to users at an appropriate level, with roles setup accordingly. Paper CRFs and consent forms will be held securely at the University Hospitals Birmingham NHS Foundation Trust and will not leave this site. The data will be securely archived on completion of the trial and will be stored as per the Data Protection Act, 2018.

### Confidentiality

Patient identifiers including date of birth, and participant names will be securely recorded within REDCap for individual participants. Postcodes will be recorded to enable estimation of multiple deprivation indices.

### Sample collection

#### Venepuncture and blood sample preparation

Blood will be collected peripherally using vacutainers. A maximum of 70mL of blood will be taken at any one time. Samples will be prepared to enable further analysis and frozen at - 80°C (this will include whole blood, peripheral blood mononuclear cells, serum, and plasma samples).

#### Adipose biopsies

Adipose will be collected via a 3cm incision in the abdominal wall through the skin to the subcutaneous layer. Forceps will be used to remove 3 small pieces of adipose tissue with an approximate total weight of 500mg. Adipose biopsies will be collected according to a Standard Operating Procedure to ensure consistent sample quality.

#### Stool sample collection

Stool samples will be collected in sterile containers and frozen at -80°C prior to analysis.

### Laboratory evaluation

In brief, we will use blood samples to assess immune subset frequencies and calculate IMM-AGE scores, T and B cell mTOR activity in response to PMA and ionomycin, T cell metabolic fitness, T cell autophagosome breakdown, epigenetic age calculated from DNA methylation, circulating soluble mediators of inflammageing, mitochondrial DNA copy number and adipose biopsies to assess number of senescent cells with SA-β-GAL and senescent burden, epigenetic age and the isolation of adipose derived stem cells to assess stem cell exhaustion. We will assess microbial composition and functional potential through metagenomics in stool to identify biomarkers of resilience.

### Storage of biological specimens

The blood and adipose samples will be stored in -80 °C freezers at the University of Birmingham.

The stool samples will be frozen at the Research Facility and kept initially in the freezers at the University of Birmingham. Once all stool samples have been collected the samples will be transferred via secure courier for storage and further analysis at the Department of Twins Research, King’s College London (our partners in the wider Wellcome Leap consortium).

Surplus samples remaining at the end of study will, with participants consent, be stored for future ethically approved research projects. Samples will be stored for up to 5 years.

## Statistical methods

### Primary outcome analysis

Paired t-tests and/or Wilcoxon signed-rank tests (dependent on the normality of the data) will be used to test for change in SA-β-Gal following the intervention. There will be subgroup analysis of each biological sex as there are known differences in senescence(95). This is a biological outcomes trial and we will include only individuals who have completed the intervention and have paired adipose tissue samples within the final analysis (Per-Protocol Approach). However, as the pharmacodynamics of the three geroprotectors are unknown, we will also conduct further subgroup analysis including individuals who started but did not complete the interventions (Intention-to-Treat).

### Secondary Outcome and Exploratory analysis

Paired t-tests and/or Wilcoxon signed-rank tests (dependent on the normality of the data) will be used to test for change in the secondary outcomes following intervention.

Additionally, we will utilise advanced machine learning techniques to identify predictors of biological effects against multiple pathways implicated with impaired resilience with ageing. Predictors will include baseline biological and clinical biomarkers. This will require an iterative process that will be modelled alongside the results of the REBOUND study. Briefly, within the REBOUND study we will use a stepwise approach of canonical correlation analysis to identify biological pathways that are implicated within clinical trajectories of impairment and non-recovery of physical and cognitive function (i.e., reduced resilience) following colorectal surgery, and LASSO and EN models to identify baseline predictors of such adverse trajectories (90). We will then identify if any of our three geroprotectors demonstrate biological effects on the implicated pathways, and assess for clinical and biological biomarkers for prediction of effectiveness at baseline.

## Oversight and monitoring

A Trial Management Group will convene approximately monthly throughout the duration of the trial. Members will consist of the Chief Investigator, local site research staff and Co-Investigators for the wider research programme. The Trial Steering Committee will be combined with the Scientific Steering Committee for the wider research programme, and we anticipate will meet three times: before recruitment begins, during recruitment and following analysis of all the data. The Trial Steering Committee will provide independent oversight. Its primary role is to monitor trial progress, ensure participant safety, maintain scientific integrity, and advise the Trial Sponsor on the trial’s conduct and continuation.

There will be no data monitoring committee given the short duration of the intervention and low risk to participants. There will be no interim analysis because it is a small trial and the risk of bias at interim analysis would be high. Audit will be conducted by the Trial Sponsor.

### Adverse Event Reporting

Adverse events (AE) will be reported by participants or other health care professionals and will be recorded from the time a participant consents to join the study until the last study visit. Participants with unresolved AE at the last study visit will be followed up until resolution or 30 days after last visit of that participant, whichever is sooner. The Chief Investigator, or delegate, will ask about the occurrence of AE and hospitalisations at the telephone calls and also at third trial visit (day 28/35). AE will be recorded on the AE Log.

Serious AE will be submitted on a Serious AE form to the Chief Investigator and study team within 24 hours of becoming aware of the Serious AE. Serious AE will be assessed for expectedness and causality by the Investigator. The Sponsor will be informed of any reportable Serious AE. The evaluation of expectedness will be made based on the knowledge of the reaction and the relevant product information

### Ancillary and Post Trial Care

Indemnity is provided by the Sponsor to cover the participants in the event of harm. Participants with unresolved AE at the last study visit will be followed up until resolution or 30 days after last visit of that participant, whichever is sooner.

## Dissemination plans

A final report of the trial will be provided to the Sponsor, Research Ethics Committee and the trial Funder within 1 year of the end of the study. The trial is registered on the ISRCTN (International Standard Randomised Controlled Trial Number) database and trial results will be made publicly available on the ISRCTN trial registry within 12 months of the end of the trial, defined as Last Patient Last Visit date.

The main trial report and any sub-studies will be published in high impact peer-reviewed gerontology journals (e.g., Age and Ageing). All funding or supporting bodies will be acknowledged within publications, but they will not have reviewed and will not have publication rights of the data from the study. The results will also be presented at academic conferences and through a series of public engagement events including webinars.

Participants enrolled in the study will be informed of the results by a written summary sent personally to them.

The anonymised participant level dataset will not be publicly available but will be available from the investigator upon reasonable request.

## Discussion

REPROGRAM and REBOUND represent an ambitious research project with the aim to drive forward understanding of the underlying biological pathways of dynamic resilience.

Running a clinical study investigating the biological pathways of dynamic resilience in a patient population simultaneously with a clinical trial investigating the ability of geroprotectors to enhance these pathways is unique and it is expected to advance our understanding significantly.

The evidence for geroprotectors has been predominantly gathered in non-human models and epidemiological studies. The results of clinical trials have thus far been disappointing(56, 57, 99). Our study provides a novel approach in studying multiple broadly implicated biological pathways, combined with deep clinical phenotyping at baseline, and utilisation of advanced machine learning techniques, whilst enabling direct linkage with a patient cohort. This approach is expected to enable a stratified medicine approach within future geroprotective trials, where interventions are carefully targeted to prevent and/or treat clinical manifestations of impaired resilience in older people in ways that ensure treatment is most likely to benefit them and not cause harm.

## Trial Status

Protocol V.3.0 (12/09/25). Recruitment began on 24/09/25 and will approximately close in June 2026.

## Data Availability

No datasets were generated or analysed during the current study. All relevant data from this study will be made available upon study completion.

## Abbreviations

AE: Adverse Event
AMPK: AMP-activated protein kinase
BIA: Bioelectrical Impedance Analysis
CRF: Case Report Form
CRP: C-Reactive Protein
DNA: Deoxyribonucleic acid
EEG: Electroencephalography
ELISA: Enzyme-linked immunosorbent assay
ISRCTN: International Standard Randomised Controlled Trial Number
MMP-2: Matrix metalloproteinase 2
MMP-9: Matrix metalloproteinase 9
MoCA: Montreal Cognitive Assessment
MR: Modified release
mTOR: Mammalian target of rapamycin
PBMC: Peripheral Blood Mononuclear Cells
REBOUND: Resilience Breakthroughs in Older people Undergoing cancer procedures
REPROGRAM: Resilience Promotion with Geroprotectors: Assessment of biological effect
SA-β-GAL: Senescence-associated beta-galactosidase
SAR: Serious adverse reaction

## Declarations

### Patient and Public Involvement

The Wellcome Leap funding application timeline did not allow for the consultation of patients or the public on trial design. However, all investigators involved in trial design had previously completed Patient and Public Engagement for similar projects and incorporated relevant information into the final trial design. There are a series of public engagement events including webinars planned for the dissemination of results. Participants enrolled in the study will be informed of the results by a written summary sent personally to them.

## Acknowledgements

Part of this study has been delivered through the National Institute for Health and Care Research (NIHR) Birmingham Biomedical Research Centre (BRC).

## Author Contributions

DW is the Chief Investigator and corresponding author. She led the protocol development, trial design and manuscript preparation. She contributed to the development of the proposal. TJ is Principal Investigator for the Wellcome Leap Funded grant and conceived the study and led the proposal. He has contributed to the protocol development and trial design. AN has contributed to trial design and manuscript preparation. JL has contributed to trial design and manuscript preparation. CW and HMM are Co-Principal Investigators and contributed to the proposal, protocol development and trial design. AA, NAD, JPM, SWJ, AM, JRH, TP, JP, TN, KT, YMS, SM, AN, NR and CS are all Co-Investigators and have contributed to the proposal, protocol development and trial design. All authors have read and approved the final manuscript.

## Funding and Sponsor Contributions

This trial is funded with a Wellcome Leap Dynamic Resilience program (co-funded by Temasek Trust). The funder and sponsor have had no role in the trial design, data collection, trial management, analysis, interpretation, report writing or decision to submit for publication.

## Availability of data and material

The anonymised participant level dataset will not be publicly available but will be available upon reasonable request. Biological samples will be stored for five years and are available for future ethically approved research projects. The Trial Steering Committee will decide upon the legitimacy of any requests for either datasets or biological samples.

## Ethics approval and consent to participate

A favourable ethical opinion has been granted from the UK Health Research Authority Research Ethics Committee (Reference approval number: 24/LO/0549). The trial has been included in the National Institute for Health and care Research Clinical Research Network (NIHR CRN) portfolio (NIHR CRN study ID: 62299). The trial Sponsor is University of Birmingham. This protocol manuscript is based on V.3.0 of the trial protocol (12/09/25). Written, informed consent will be obtained from all participants.

## Consent for publication

Not applicable.

